# IL-33 expression in response to SARS-CoV-2 correlates with seropositivity in COVID-19 convalescent individuals

**DOI:** 10.1101/2020.07.09.20148056

**Authors:** Michal A. Stanczak, David E. Sanin, Petya Apostolova, Gabriele Nerz, Dimitrios Lampaki, Maike Hofmann, Daniel Steinmann, Robert Thimme, Gerhard Mittler, Cornelius F. Waller, Edward J. Pearce, Erika L. Pearce

**Author notes:** equal contribution (Michal A. Stanczak, David E. Sanin, Petya Apostolova, Cornelius F. Waller, Edward J. Pearce, Erika L. Pearce).

## Abstract

Our understanding of severe acute respiratory syndrome coronavirus 2 (SARS-CoV-2) is still developing. We investigated seroprevalence and immune responses in subjects professionally exposed to SARS-CoV-2 and their family members (155 individuals; ages 5–79 years). Seropositivity for SARS-CoV-2 spike glycoprotein aligned with PCR results that confirmed previous infection. Anti-spike IgG titers remained high 60 days post-infection and did not associate with symptoms, but spike-specific IgM did associate with malaise and fever. We found limited household transmission, with children of infected individuals seldomly seropositive, highlighting professional exposure as the dominant route of infection in our cohort. We analyzed PBMCs from a subset of seropositive and seronegative adults. TLR7 agonist-activation revealed an increased population of IL-6^+^TNF^-^IL-1β^+^ monocytes, while SARS-CoV-2 peptide stimulation elicited IL-33, IL-6, IFNa2, and IL-23 expression in seropositive individuals. IL-33 correlated with CD4^+^ T cell activation in PBMCs from convalescent subjects, and was likely due to T cell-mediated effects on IL-33-producing cells. IL-33 is associated with pulmonary infection and chronic diseases like asthma and COPD, but its role in COVID-19 is unknown. Analysis of published scRNAseq data of bronchoalveolar lavage fluid (BALF) from patients with mild to severe COVID-19 revealed a population of IL-33-producing cells that increases with disease. Together these findings show that IL-33 production is linked to SARS-CoV-2 infection and warrant further investigation of IL-33 in COVID-19 pathogenesis and immunity.

## MAIN

We took the opportunity to study a cohort of individuals with a known professional exposure, within a relatively small timeframe (∼2–3 months), as the pandemic was developing. We assessed viral transmission among these individuals and their family members. Specifically, we used an ELISA to measure IgG and IgM titers against the SARS-CoV-2 spike glycoprotein^1^. Our study population included 155 subjects who either tested positive for SARS-CoV-2 via PCR or were in contact with SARS-CoV-2-infected individuals. Demographic and disease characteristics of the study population are indicated in Supplementary Table 1. Median age was 36 years and 98 (63.2%) were females; 99 (64.5%) had professional exposure to the virus, 44 (28.4%) were exposed within their household and 12 subjects had either multiple or different/unknown exposure sources. Symptoms of upper respiratory tract infection within the last 16 weeks prior to blood collection were reported by 97 (62.6%) individuals. SARS-CoV-2 PCR testing was performed in 87 individuals, with 47 (30.3%) having a positive PCR result. We evaluated the presence and abundance of antibodies against the spike protein of SARS-CoV-2 in the serum of the study population, as well as in 16 historical sera collected prior to the onset of the COVID-19 pandemic. We found highest antibody titers among individuals with a positive SARS-CoV-2 PCR (**Fig. 1a**), as well as a few cases where subjects with a negative PCR result possessed antibodies against the viral protein (9/40). As the date of the PCR and the collection of serum samples were separated by a median time of 49 days, it is possible that these individuals became infected after a PCR was carried out. Likewise, we discovered few cases (3/47) where a positive PCR result did not associate with elevated anti-spike IgG titers.

**Figure 1.**
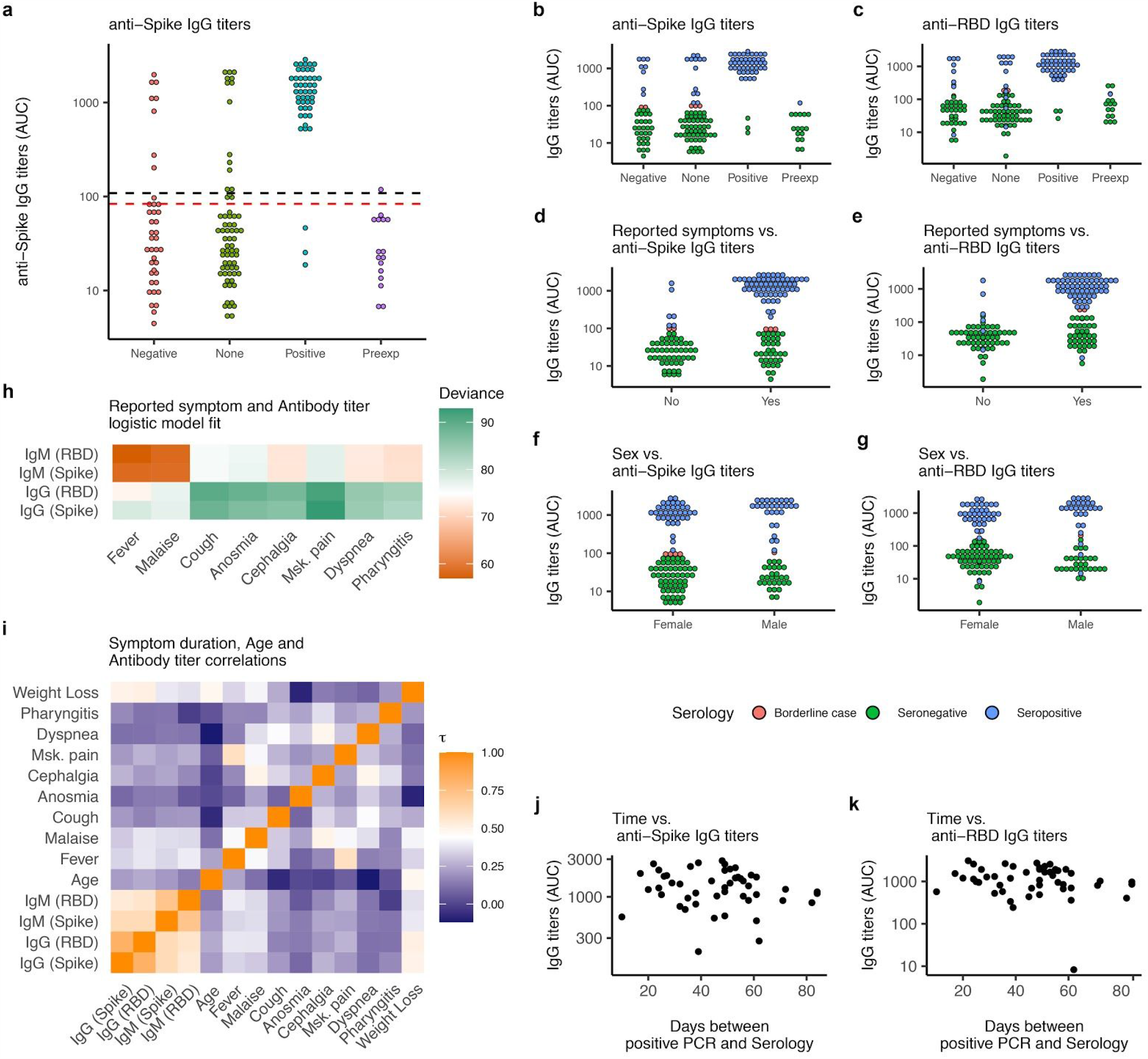
Serological characterization of study population. **a**, Anti-spike IgG titers shown as area under titration curve (AUC) in sera from investigated subjects grouped by SARS-CoV-2 PCR status (Negative, None, Positive) plus pre-pandemic historical sera (Preexp). Dotted lines illustrate estimated thresholds above which subjects were classified as “Borderline case” (mean + 2xSD – red) or “Seropositive” (mean + 3xSD – black). Mean and SD were calculated for all IgG titers below maximum value measured from Preexp sera. **b–g**, Anti-spike (b, d, f) and anti-RBD (c, e, g) IgG titers shown as AUC for investigated subjects grouped by SARS-CoV-2 PCR status (b–c), reported symptoms (d–e) and sex (f–g). Subjects are colored by serology results. **h**, Detected antibody titers were used to fit logistic regression models to symptom reporting by seropositive subjects. Deviance from logistic model fitting is shown as a measure of model accuracy. **i**, Detected antibody titers from seropositive subjects were correlated to each other, symptom duration and subject age using a Kendall rank correlation. A heatmap of the Kendall rank correlation coefficient (*τ*) is shown for each pair of variables. **j–k**, Anti-spike (j) and anti-RBD (k) IgG titers shown as a function of time for seropositive subjects. Approximated time of infection was taken as day of positive SARS-CoV-2 PCR status.

We used the signal measured in pre-exposure sera to set a threshold to define seropositive and borderline cases (**Fig. 1a** – black and red dashed lines respectively). This resulted in 5 borderline cases and 65 seropositive individuals. We then probed collected sera for IgG and IgM antibodies against the spike protein and its receptor binding domain (RBD) finding a robust correlation between these measurements (**Fig. 1b–c, i** and **Extended Data Fig. 1a–e**). Most seropositive subjects reported experiencing symptoms (**Fig. 1d**). We discovered only 6 asymptomatic individuals out of 65 seropositive subjects (2/8 borderline cases) who nonetheless had high anti-spike IgG titers (**Fig. 1d**) although anti-RBD IgG titers were appreciatively lower (**Fig. 1e**). We found no association between sex and antibody titers (**Fig. 1f–g**). To explore possible links between clinical features and antibody titers, we probed our data with two complementary strategies. First, we fitted logistic regression models to reported symptoms among seropositive subjects (**Fig. 1h** and **Extended Data Fig. 1f–h**), setting the symptom report (yes = 1, no = 0) as the outcome predicted by the antibody titer and calculating the deviance of the model as a measure of accuracy. Within our cohort, elevated IgM levels against spike and RBD were reasonable predictors of experiencing malaise and fever (**Fig. 1h** and **Extended Data Fig. 1f–g**), but not for other symptoms (**Extended Data Fig. 1h**). These results might suggest that people with strong immune responses against the virus feel sicker during acute infection, but also have persistent IgM antibodies as a result of this stronger antiviral response. Next, we calculated the Kendall’s rank correlation coefficient (*τ*) between symptom duration, subject age, and antibody titers in pairs (**Fig. 1i**). Beyond the strong correlation between IgG and IgM titers for both spike and RBD, we found modest to no correlation among clinical features. Finally, we observed no appreciable drop in antibody titers in the study population, even 60 days after active infection (**Fig. 1j–k** and **Extended Data Fig. 1i–j**).

We sought to visualize viral transmission within our study population by generating an undirected network of interactions based on subject exposure history (**Fig. 2a**). We limited our analysis to subjects with professional exposure plus all their adjacent nodes. The dense and highly connected center of the network corresponds to professional exposure, while family groups within households are arranged in the periphery (**Fig. 2a**). We observed that a large subset of our study population was linked by an episode of professional exposure resulting in infection, however the extent to which this event caused viral spread to subjects’ households was limited. In fact, upon closer analysis of household groupings (**Fig. 2b**) we found low levels of transmission. Indeed, when we measured the assortativity of these household network groups, as a proxy for viral transmission rate, we confirmed that for a network of the size and distribution of our study the observed assortativity with respect to serology status and IgG titers was remarkably low (**Fig. 2c** - observed assortativity in red) indicating a low transmission rate. Children (thicker edged nodes) in particular were seldomly positive in our serology results (3/12), despite coming into contact with infected individuals with high anti-spike IgG titres.

**Figure 2.**
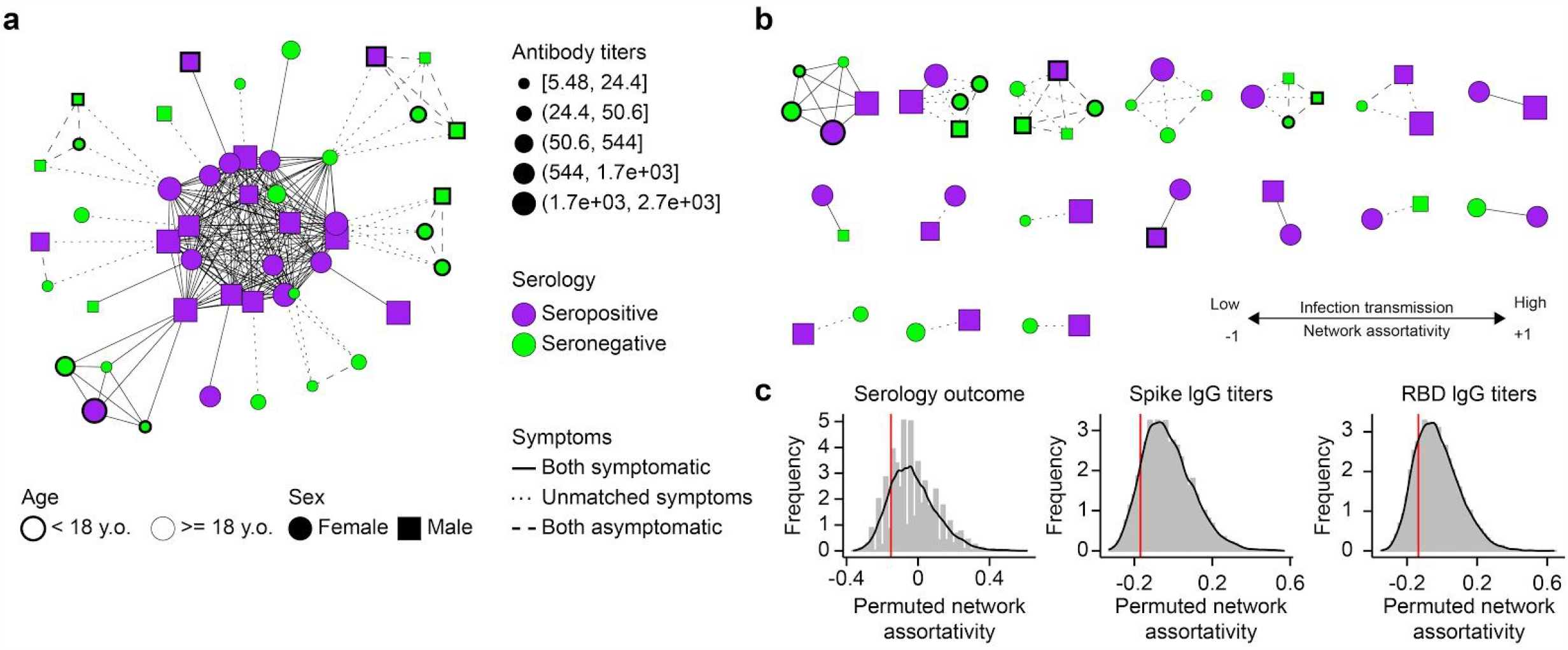
Professional exposure, rather than household exposure, dominates SARS-CoV-2 spread. **a**, Subjects with known professional exposure or sharing a household with subjects with known professional exposure to SARS-CoV-2 were organized in an undirected network. Edges connect individuals exposed to one another in a professional (central arrangement) or household setting. Individual’s anti-spike IgG titers (symbol size), serology outcome (symbol color), age (symbol border) and sex (Symbol shape) are illustrated in the network. Connecting edges highlight matching symptoms between individuals. **b**, As in a but connections limited to household exposure (i.e. showing family groups). **c**, Network assortativity, used as an indication of infection transmission rate, was calculated based on the indicated parameters for the network in b (red line) and also after parameter randomization over 10000 permutations of the network (shown as a distribution).

SARS-CoV-2 represents a unique public health challenge in modern times, with the possibility of subsequent waves of infection being an active area of research. While antibodies can neutralize the virus and are therefore likely to be important for protective immunity^2^, it is probable that the pathologic effects of SARS-CoV-2 infection are cell-mediated. The development of IgG antibodies during infection is indication that a CD4^+^ T follicular helper cell response is mounted against this virus^3,4^, and recent work has indicated that both CD4^+^ and CD8^+^ T cells capable of making IFN-γ and TNF are present within the blood of convalescent patients^4–6^. We therefore decided to look broadly at cellular immune responses in convalescent patients. We focused on both SARS-CoV-2 spike protein-specific T cell responses and innate monocyte responses, and measured cytokines known to be produced by both adaptive and innate immune cells. We did this for a subset of 20 convalescent individuals in our cohort, comparing them to 20 seronegative subjects in our study (Supp. Table 2). We purified PBMCs from these individuals and examined their composition (**Fig. 3a–d**) and activation following stimulation (**Fig. 3a, e–h**), correlating these results to antibody titers (**Fig. 3** in bold) using conventional markers (**Extended Data Fig. 2**) and 2 different stimuli (**Fig. 3a**). Overall, we did not detect a strong association between the cellular composition, immune cell activation, time from active infection, and antibody titers, (**Fig. 3** and **Extended Data Fig. 3**) with the latter more closely associated with subject age than any other measured parameter (**Fig. 3b–f**). We also observed a bimodal distribution of seropositive subjects, with the studied population clustering in 2 separate groups (**Fig. 3**) indicating an underlying complexity in our cohort. However, upon examination of TLR7-driven classical monocyte activation (**Fig. 3g**), we discovered that the percentage of IL-6^+^TNF^-^IL-1β^+^ monocytes clustered closely with IgG titers. Moreover, SARS-CoV-2 peptide-induced production of IL-33, IL-6, IFN-α2 and IL-23 in these cultures (**Fig. 3h**) also clustered together with IgG titers. Critically, IL-33 production was more closely related to IgG titers than any other measured parameter (**Extended Data Fig. 3**), including subject age.

**Figure 3.**
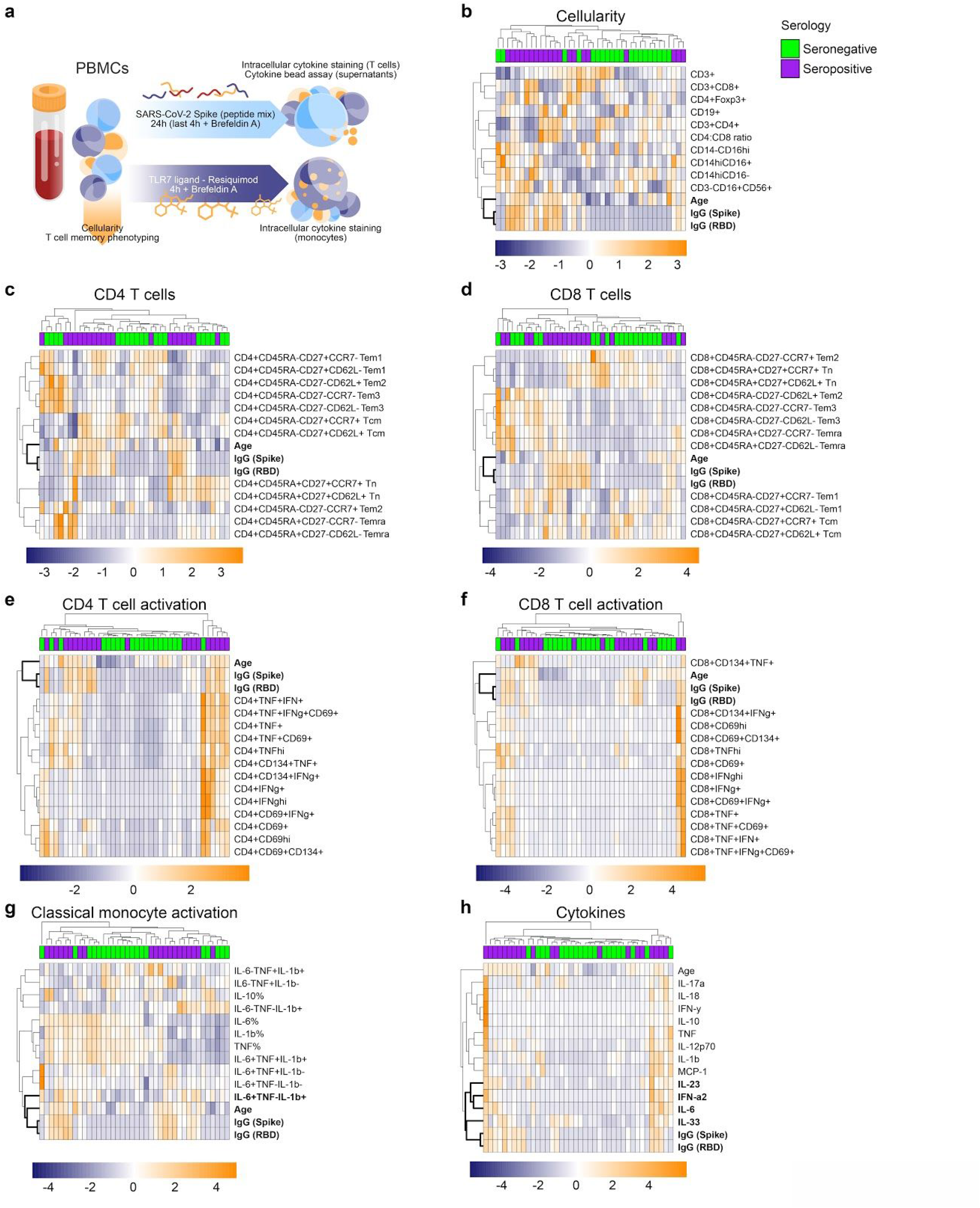
PBMCs from convalescent and non-infected subjects differ only in innate cytokine production profile. **a–h**, PBMCs in blood collected from a subset of 20 seropositive and 20 seronegative subjects were isolated, analyzed via flow cytometry for cellular composition (b–d) and stimulated with a SARS-CoV-2 spike peptide mix or vehicle control (e–f, h), or with TLR7 agonist Resiquimod or vehicle control (g) for indicated times (a). Activation of CD3^+^CD4^+^ (e), CD3^+^CD8^+^ (f) and CD14^hi^CD16^-^ cells (g) plus cytokine secretion into culture media (h) were measured via flow cytometry. Control corrected (treatment - control) scaled data 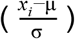 was then visualized as heatmaps. Scaled anti-spike and anti-RBD IgG titers plus subject age were included in results. Data was clustered based on Euclidean distances between features (rows) or subjects (columns), with clusters indicated with dendrograms. Serology results are included for each subject (seropositive – purple; seronegative – green). Closest associations to antibody titers are highlighted in bold letters.

While resistance to infection is likely to involve cytokines such as IFN-γ produced by T cells, many of the symptoms associated with severe COVID-19 appear to be linked to excessive production of innate cytokines such as IL-6 and IL-1β^7^. Consequently, we felt that the results highlighting IL-33, IL-6, IFN-α2, IL-23 and IL-1β production (**Fig. 3**) could be important. In line with our clustering analysis, we found that all highlighted features were in fact significantly increased in seropositive subjects (**Fig. 4a–e**). We reasoned that the observed cytokine production (**Fig. 4a–d**) should be a result of T cell activation, as our experimental strategy favours this mechanism. We found that this could be the case for IL-33, which had a strong positive correlation with CD69 expression in CD4^+^ T cells (**Fig. 4f**). However, this was not the case for other parameters (**Fig. 4g–i**) and not unexpectedly for the percentage of IL-6^+^TNF^-^IL-1β^+^ monocytes (**Fig. 4j**).

**Figure 4.**
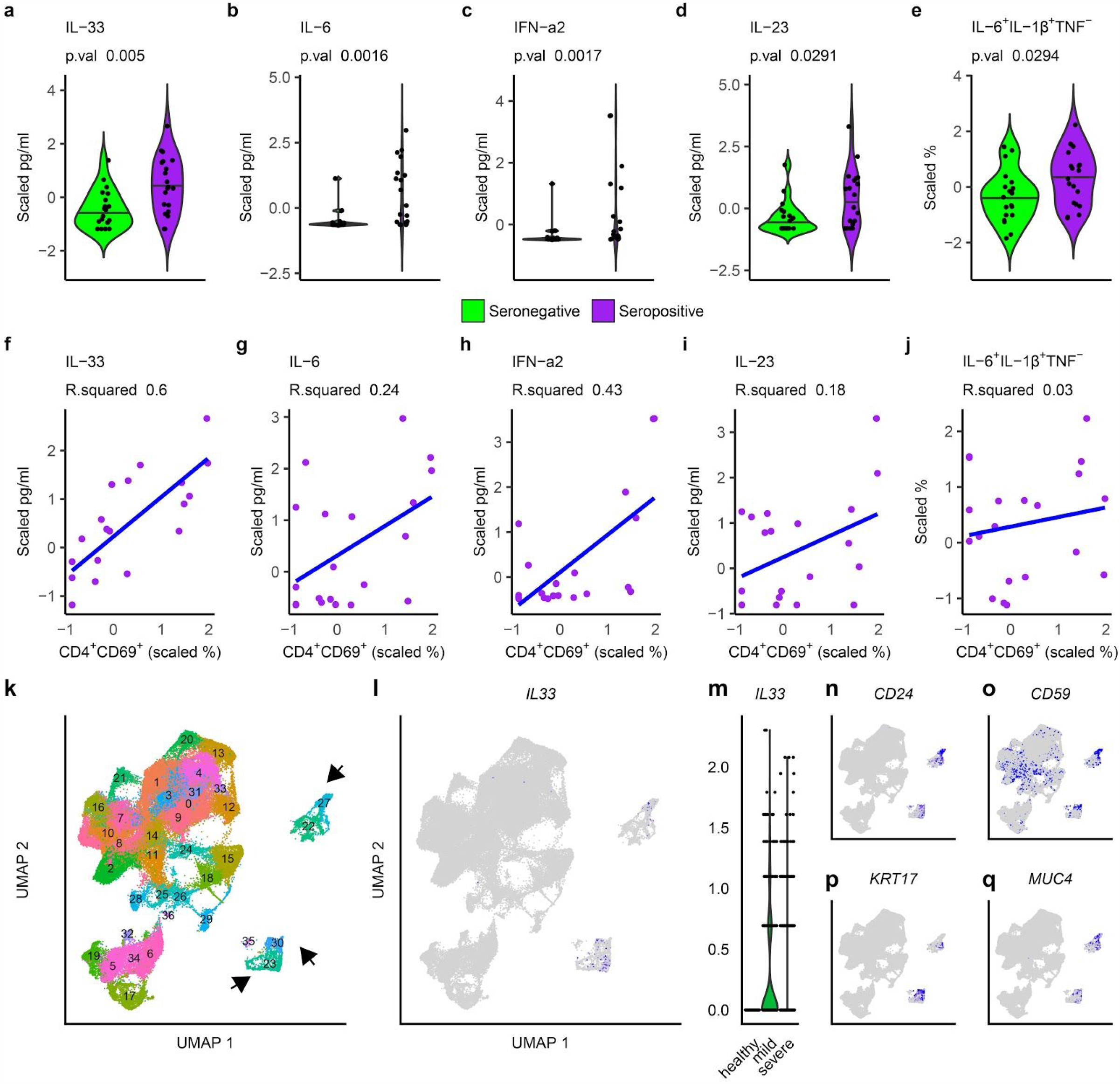
IL-33 production correlates with T cell activation and disease severity in SARS-CoV-2 infected subjects. **a–e**, Cytokine production measured in the media of cultured PBMCs (a–d) or via intracellular staining of CD14^hi^CD16^-^ monocytes from seropositive (purple) and seronegative (green) subjects is shown as scaled values. Two tailed Mann-Whitney U tests were calculated and resulting p value is reported. **f–j**, Scaled cytokine production versus corresponding CD4 T cell activation (CD4^+^CD69^+^) in PBMC cultures of seropositive individuals is shown. Linear regression curves fitted to these data (blue) alongside R^2^ values are provided. **k–q**, Single cell RNA sequencing data from the bronchoalveolar lavage fluid of 3 healthy individual compared to 9 SARS-CoV-2 infected subjects with different disease severities (3 mild; 6 severe) was retrieved from a public database (GSE145926), grouped into cell clusters (k) and analyzed for the expression of IL-33 (l). **m**, IL-33 expression in clusters 23, 27 and 30 presented as a violin plot grouped by disease status. **n–q**, Key surface (n–o) and lineage specific (p–q) genes expressed in IL-33 producing cells.

To our knowledge, there are no reports associating IL-33 with COVID-19 and given the strong correlation we observed in our data we decided to explore this result in more detail. To accomplish this, we retrieved single cell RNA sequencing data from a recent publication^8^ and processed it to interrogate *IL33* expression in the setting of SARS-CoV-2 pulmonary disease (**Fig. 4k–q** and **Extended Data Fig. 4**). These data were generated from cells in BALF of 3 healthy donors and 9 acutely SARS-CoV-2 infected patients, 3 of whom displayed mild symptoms, while the remaining 6 were classed as severely ill (**Extended Data Fig. 4**). In these data, we identified 2 potential sources of IL-33 production coming from 3 distinct cell clusters (**Fig. 4k–l**). Strikingly, *IL33* expression was confined to SARS-CoV-2 infected patients (**Fig. 4m**), concomitant with the increased appearance of the *IL33*^+^ cell clusters as a result of disease (**Extended Data Fig. 4**). *IL33*^+^ cell clusters were most likely stromal cells (**Fig. 4n–q**) and are unlikely to be responsible for the IL-33 production we observed in PBMC cultures. However, these clinical data further implicate IL-33 in COVID-19 immunobiology.

Here we show that after the resolution of infection, an innate immune cytokine signature persists in PBMCs of convalescent subjects, characterized by induction of IL-33 in response to T cell activation with 14-mer peptides, a peptide length suggestive of CD4^+^ T cell activation^9^. However, the identity and mechanism of activation of the IL-33-producing cells in the PBMCs from convalescent individuals remain unclear at this time. IL-33 was initially described as a cytokine produced as an alarmin by epithelial cells to induce type 2 immune responses^10^, and of interest in this regard, there are reports of eosinophilia, a mark of type 2 immunity, in COVID-19 patients^11^. IL-33 is also linked to lung injury, pulmonary viral infections, and chronic lung diseases^12^. COVID-19 can cause complications such as pneumonia and acute respiratory distress syndrome, as well as invoke lasting damage to lungs, including lung remodeling and fibrosis, which are consistent with known roles of IL-33^13^. These observations suggest that IL-33 could participate in the pathogenesis of COVID-19. On the other hand, IL-33 has also been reported to promote antiviral cytotoxic T cell responses^14–16^ and higher antibody production^17,18^. Thus the presence of a persistent population of cells capable of making IL-33 in response to T cell activation may confer an advantage in the case of secondary exposure. Further work on IL-33 will be needed to elucidate the role of this cytokine in COVID-19.

## Data Availability

This study does not contain large publicly available datasets.

## METHODS

### Human subjects

All procedures involving human subjects were approved by the Ethics Committee of the Medical Center - University of Freiburg (305/20). The study was registered at the German Clinical Trial Register (DRKS00022292). Pre-exposure donor serum samples (n=16) had been collected, aliquoted and stored at the Medical Center - University of Freiburg prior to June 1st, 2019. These individuals were considered to be pre-exposure controls since their biological material had been stored prior to the emergence of SARS-CoV-2. This cohort included 50% males (n=8) and 50% females. Median age was 51 years with a range of 39–77 years.

SARS-CoV-2-exposed subjects (n=155) were recruited with a standardized procedure based on assessment of inclusion and exclusion criteria. Inclusion criteria were contact to SARS-CoV-2-infected individuals, age of at least 5 years and the ability to provide a written informed consent. The exclusion criterium was the presence of symptoms of an acute SARS-CoV-2 infection (fever, cough, malaise) within the last 14 days prior to blood collection. Study participants gave their written informed consent prior to study enrollment. For subjects <18 years of age, written informed consent was provided by one parent. Collection of biological material was performed at the Medical Center - University of Freiburg. Serum and peripheral blood mononuclear cells (PBMCs) were collected at one or two time points. All samples were de-identified and analyzed in a pseudonymized manner. Information on demographic characteristics and disease course was provided by the subjects using a standardized questionnaire filled at the time point of enrollment. Information on age, sex, PCR testing for SARS-CoV-2, symptoms, symptom duration, treatment and exposure were analyzed. The cohort included 98 females (63.2%) and 57 males (36.8%). Median age was 36 years with a range of 5–79 years. Further characteristics are included in Supplementary Table 1.

### Serum asservation and ELISA

Serum was collected in tubes containing clot activators (7.5 ml) and stored at room temperature prior to processing. Tubes were centrifuged at 2500 g for 10 min and serum was aliquoted and stored at −20 °C.

### Serology

Collected serum was analyzed for the presence of antibodies against SARS-CoV-2 proteins by adapting a serology assay developed in the laboratory of Dr. Florian Krammer^1^, with some alterations to the protocol. Recombinant spike protein and its receptor binding domain (RBD) were expressed from mammalian expression plasmids kindly donated by Dr. Krammer^1^. Recombinant proteins were produced employing the Expi293 Expression System (Thermo) following the manufacturer’s recommendations. Briefly, cells were grown to exponential phase in serum-free optimized media, then transfected with plasmid-lipid complexes prepared with Expifectamine. 20 h later media were supplemented with expression enhancers and finally media containing recombinant proteins was harvested 72 h later. Proteins were purified by passing media through Ni-NTA Sepharose columns. Purity and identity of recombinant proteins was established by polyacrylamide gel electrophoresis and mass spectrometry.

96-well flat bottom plates (Corning) were coated with 2 µg/ml of recombinant proteins overnight at 4 °C. Blocking was performed with 200 µl/well PBS containing 0.05% Tween-20 (PBS/T) and 3% milk for 4 h. Serum samples were diluted 1:50, 1:200, 1:800 and 1:3200 performing serial dilutions in PBS/T containing 1% milk. Serum samples were incubated for 2 h at room temperature. Plates were washed four times with PBS/T and incubated with an anti-human IgG or IgM secondary antibody (dilution 1:3000, Sigma or Thermo Fisher Scientific) for 1 h at room temperature. Plates were washed four times with PBS/T and developed using a TMB substrate kit (Biolegend) for 5 min at room temperature. Colorimetric reaction was stopped by the addition of 2N H_2_SO_4_. Internal positive and negative standards were included on each plate to allow for data normalization between different plates. Absorbance at 450 nm was measured using a TriStar microplate reader (Berthold Technologies).

### PBMC collection

Peripheral blood from 20 seronegative and 20 seropositive subjects (based on anti-spike IgG ELISA) was collected in EDTA-coated tubes and stored at 4 °C prior to processing. Blood was diluted 1:1 with PBS containing 2% FCS and 1 mM EDTA. Peripheral blood mononuclear cells (PBMCs) were isolated using a density gradient method. Lymphoprep solution (Stem Cell) was pipetted to the bottom of SepMate-50 tubes (Stem Cell), and diluted blood carefully layered on top. After 10 min centrifugation at 1200 x g, the upper phase containing mononuclear cells was transferred to a new tube. PBMCs were washed twice with PBS with 2% FCS and 1 mM EDTA. After counting, cells were aliquoted, frozen in FCS containing 10% DMSO and stored at −80 °C.

### SARS-CoV-2 peptide stimulation

PBMCs were thawed and rested in RPMI containing 10% FCS, 100 U/ml penicillin/streptomycin, 4 mM glutamine and 55 µM 2-mercaptoethanol for 2 h. 4×10^5^ PBMCs were incubated with 2 µg/ml SARS-CoV-2 spike PepMix (JPT) or DMSO control in the presence of 2 µg/ml anti-CD28 and 50 U/ml hIL-2. The PepMix includes a total of 315 14-mer peptides covering the whole spike protein. Incubation was performed for 24 h with addition of 5 µg/ml Brefeldin A (Biolegend) for the last 4 h to block cytokine secretion. DMSO controls served as negative controls.

### TLR7 agonist stimulation

2×10^5^ PBMCs were cultured in RPMI containing 10% FCS, 100 U/ml penicillin/streptomycin and 4 mM glutamine supplemented with 5 µg/ml Brefeldin A and 5 µg/ml Resiquimod (Invivogen) or water control for 4 h.

### Flow cytometry

Prior to cell surface staining, PBMCs were incubated with purified NA/LE Human BD Fc block (BD, dilution 1:300) and Live/Dead Fixable Aqua or Live/Dead Fixable Blue (both Thermo Fisher, dilution 1:1000) in FACS buffer (2% FCS and 1 mM EDTA in PBS) for 20 min at 4 °C. Cells were then washed and incubated with cell surface antibody cocktails for 30 min at 4 °C. Cells were washed with FACS buffer and fixed using BD Cytofix Fixation Buffer (for intracellular cytokine staining) or eBioscience Fixation/Permeabilization kit (for Foxp3 staining) for 20 min at room temperature. For intracellular staining assays, cells were washed twice with 1x BD Perm/Wash buffer or 1x eBioscience Perm/Wash buffer prior to incubation with intracellular antibody cocktails for 45 min at room temperature. Cells were washed twice with 1x BD Perm/Wash buffer or 1x eBioscience Perm/Wash buffer and resuspended in FACS buffer. Flow cytometry data were acquired on a BD LSRFortessa or BD FACSymphony flow cytometer.

### Cytokine measurement

Supernatants from PBMCs cultured with SARS-CoV-2 PepMix or DMSO control as described above were collected and stored at −20 °C. Cytokines in culture media were detected using the Legendplex™ Human Inflammation Panel 1 (Biolegend, Cat# 740809) following the manufacturer’s instructions. This kit allows for simultaneous measurement of IL-1β, IFNα2, IFNγ, TNF, MCP-1, IL-6, IL-8, IL-10, IL-12p70, IL-17A, IL-18, IL-23 and IL-33. Briefly, standards and cell culture supernatants were incubated on a 96-well V-bottom plate with capture beads for 2 h. The plate was washed and biotinylated detection antibodies were added to each well for 1 h, followed by Streptavidin-phycoerythrin (PE) addition for 30 min. Samples were washed and resuspended in FACS buffer. All steps were carried out at room temperature. Results were obtained using a BD LSRFortessa flow cytometer. Data analysis was performed in FlowJo. Cytokines were identified based on the size and internal dye of the beads. Cytokine concentration in samples was derived from the geometric mean fluorescence intensity for PE interpolated from standard curves calculated from 8 standard controls measured in duplicate.

### Data analysis and Statistics

Flow cytometry data were analyzed using FlowJo version 10.6. Clinical data, ELISA readings, flow cytometry results and single cell RNA sequencing data were processed and analyzed using R (Lucent Technologies), which was also employed to generate graphs.

#### Antibody titers

Background measurements were subtracted from absorbance readings, which were then batch corrected based on internal standards included on each plate. Batch and background corrected data were then used to calculate the area under the dilution curve (AUC) for each subject. To calculate a seropositivity threshold, a subset of AUC values was extracted, composed of all pre-pandemic sera as well as those subjects with an AUC lower than the maximum obtained from pre-exposure sera and with no SARS-CoV-2 PCR or with a negative PCR result. Borderline cases were defined as being 2 standard deviations away from the mean AUC from this sample subset, while AUC results at least 3 standard deviations away from the mean were taken as seropositive. All other values were defined as seronegative.

#### Symptoms, age and titer associations

The relationship between antibody titers, subject age and reported symptoms were explored in two parallel ways. First, antibody titers were used to fit a logistic regression model capable of predicting the presence of reported symptoms (yes = 1, no = 0). The deviance of each model was used to estimate the quality of the fit. Next, antibody titers, subject age, and symptom duration in days were correlated with one another using Kendall’s rank correlation and reporting *τ* as the correlation coefficient. Only complete pairs of observations were used in correlations and all data was scaled by subtracting the mean of each feature and dividing by the corresponding standard deviation.

#### Network analysis

An adjacency matrix was generated for the network of subject interactions based on reported exposures. Subjects with suspected professional exposure to SARS-CoV-2, who shared a workplace, were connected to each other. Subjects belonging to the households of professionally exposed individuals were added to the network, with interactions within households included. Only subjects with known professional exposure and with at least one household connection were included in this analysis. To estimate viral transmission within households, a network was constructed exclusively with household interactions and later the assortativity of the network was estimated with regards to serological results and antibody titers. Observed assortativities were compared to those calculated based on 10000 permutations of used values over the same network architecture. Network visualization, analysis and construction was performed with igraph v.1.2.5.

#### Immunophenotyping clustering and regression

Flow cytometry results were collected and stimulation controls were subtracted from values obtained for treatments. Corrected values were subsequently scaled and clustered based on Euclidean distances to antibody titers and subject age, which were similarly scaled. Heatmaps and clustering were obtained by using pheatmap v.1.0.12. Selected features were correlated with one another using a linear regression model (y ∼ x) and reporting R squared as an indication of the quality of the fit.

#### Single cell RNA sequencing analysis

The transcriptional profiles of ∼90000 single cells in bronchoalveolar lavage fluid from 3 healthy individuals and 9 SARS-CoV-2 infected patients (3 with mild and 6 with severe disease) were retrieved from a publicly available repository (GSE145926). Matrices with raw unfiltered read counts for detected features in barcoded cells were downloaded for each subject and subsequently processed, analyzed and visualized in R using Seurat v. 3^19^, removing cells with a high percentage of mitochondrial RNA over total RNA (> 25%) and with less than 200 detected features, normalizing gene expression data by applying regularized negative binomial regression^20^ and using a Uniform Manifold Approximation and Projection (UMAP)^21^ as a dimensionality reduction approach. Differentially expressed genes within each cluster and across conditions were determined with Seurat as those with a greater than 1.2 fold change and an adjusted p value of less than 0.05.

## ACKNOWLEDGEMENTS

The authors would like to thank Yvonne Neumann, Johan Fridén, Florian Krammer, Mauro Corrado, Asifa Akhtar, Bertram Bengsch, Martin Schwemmle, Evelyn Pearce, Constanze Waller, Fabian Hässler, and Dan Puleston for their contributions to the study.

## AUTHOR CONTRIBUTIONS

M.A.S., D.E.S., P.A., C.F.W., E.J.P. and E.L.P designed the research, provided conceptual input and analyzed the data. M.A.S., D.E.S., P.A., G.N., D.L. and G.M. performed the experiments. P.A., M.H., D.S., R.T. and C.F.W. recruited participants and collected samples plus clinical information. M.A.S., D.E.S., P.A., E.J.P. and E.L.P wrote the manuscript.

## FUNDING

This study was supported by the Max Planck Society, the German Research Foundation (DFG) Leibniz Prize (E.L.P.), and the DFG under Germany’s Excellence Strategy (CIBSS EXC-2189 Project ID 390939984). M.A.S. received funding from the Swiss National Science Foundation.

Correspondence and requests for materials should be addressed to E.L.P., E.J.P. or C.F.W.

---

**Extended Data Figure 1.**
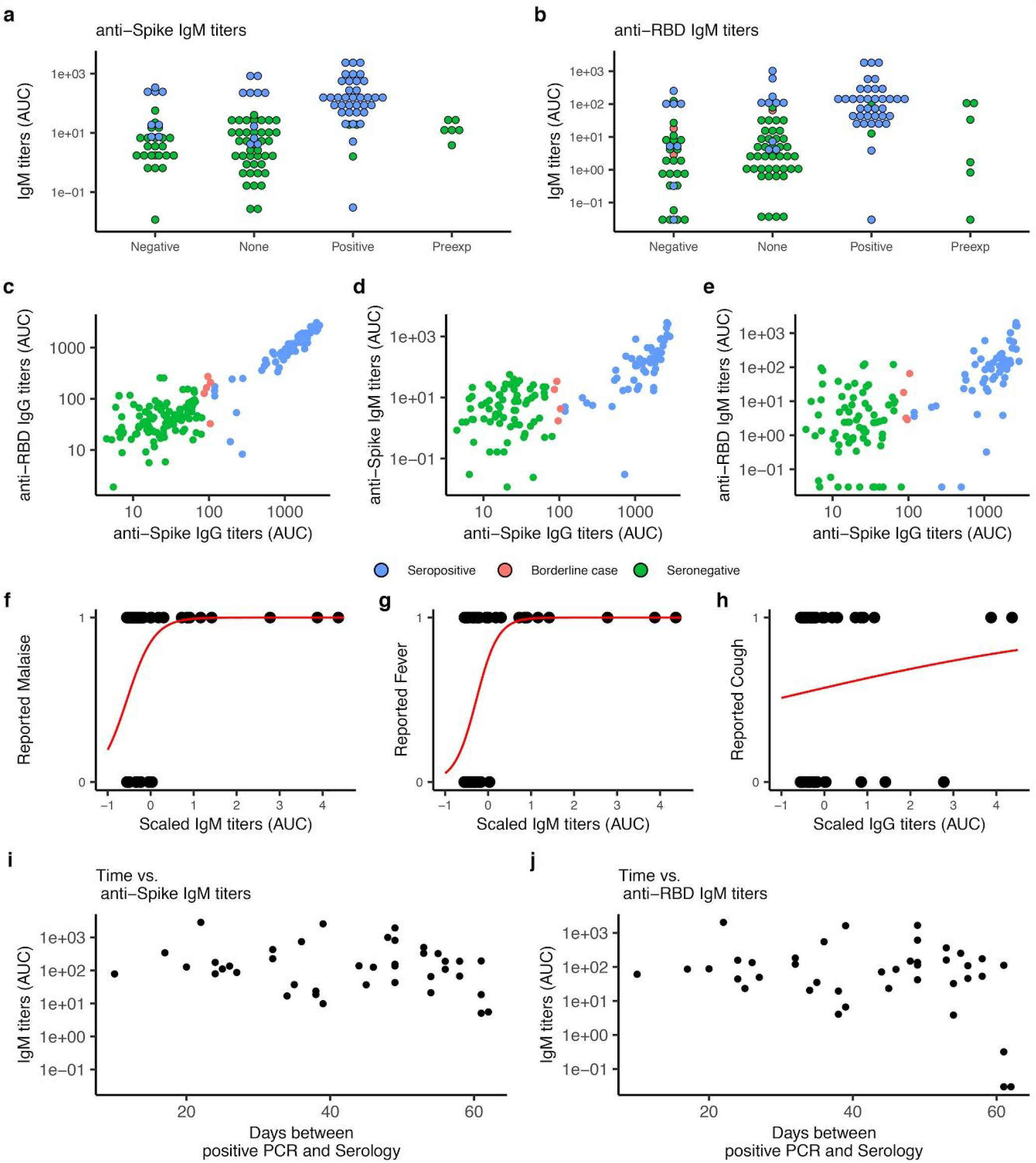
Related to Figure 1: Serological characterization of study population. **a–b**, Anti-spike (a) and anti-RBD (b) IgM titers shown as AUC for investigated subjects grouped by SARS-CoV-2 PCR status. Subjects are colored by serology results. **c–e**, Indicated antibody titers are shown versus anti-spike IgG titers, with subjects colored by serology results. **f–h**, Detected antibody titers were used to fit logistic regression models to symptom reporting by seropositive subjects. Fitted models (red lines) with low deviance (Malaise – f and Fever – g) as well as with high deviance (Cough – h) are shown. **i–j**, Anti-spike (j) and anti-RBD (k) IgM titers shown as a function of time for seropositive subjects. Approximated time of infection was taken as day of positive SARS-CoV-2 PCR status.

**Extended Data Figure 2.**
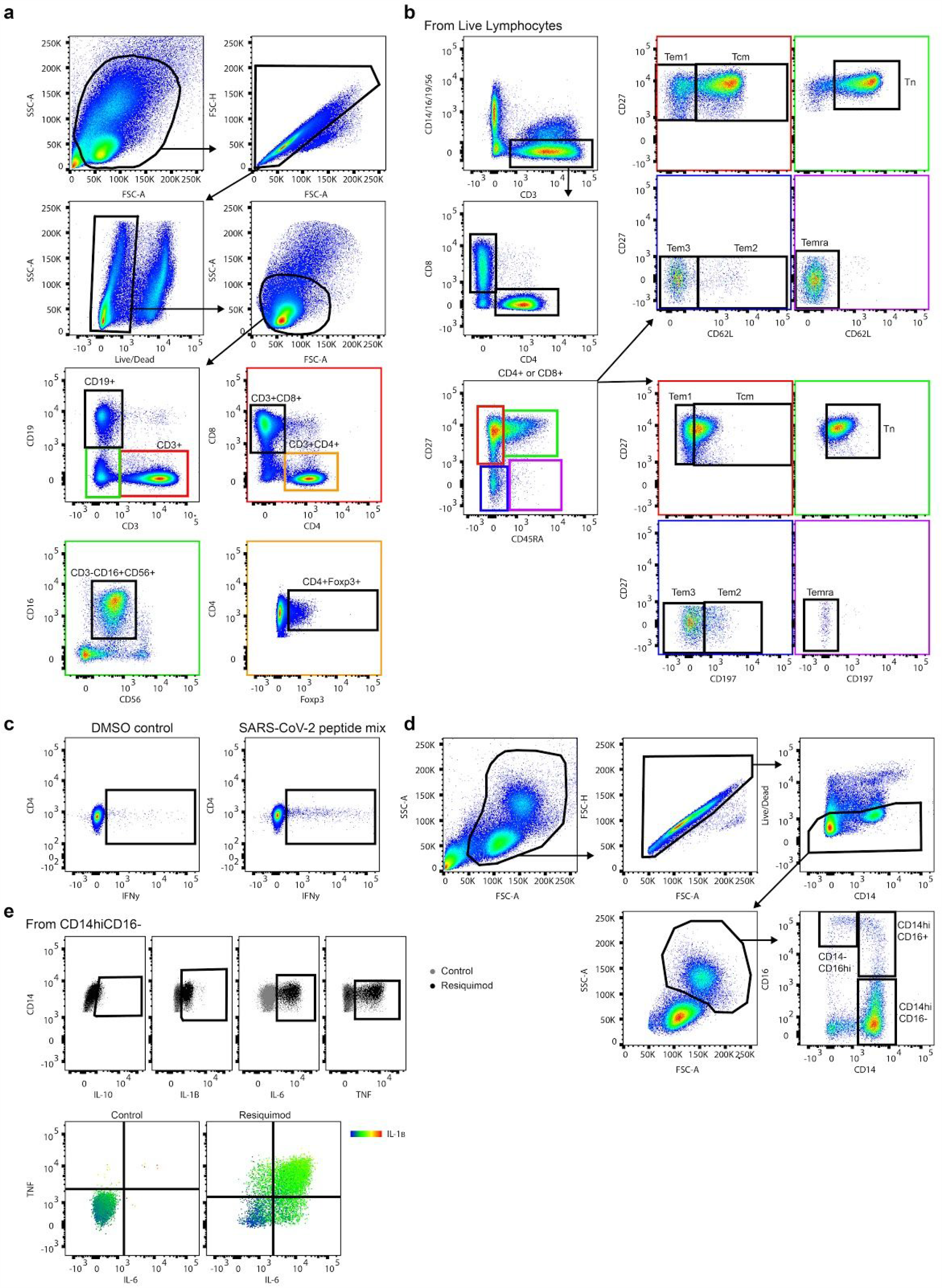
Related to Figure 3: PBMCs from convalescent and non-infected subjects differ only in innate cytokine production profile. **a–e**, Gating strategies for PBMCs shown as representative flow cytometry dot plots for lymphocytes (a), CD4^+^ and CD8^+^ memory cell populations (b), lymphocyte cytokine production (c), monocyte populations (d) and monocyte cytokine production (e). Control and treatments results are shown for stimulation experiments (c, e).

**Extended Data Figure 3.**
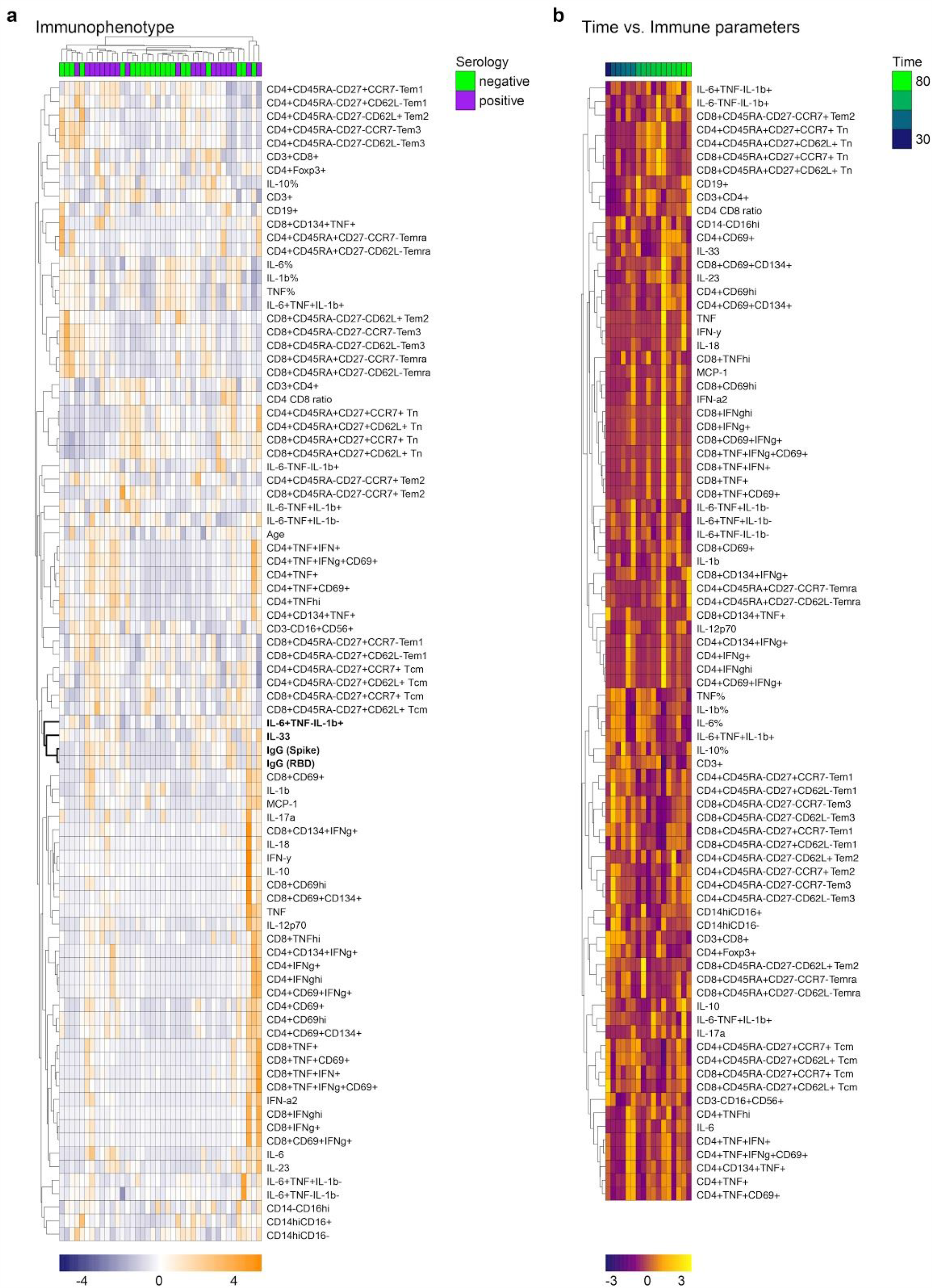
Related to Figure 3: PBMCs from convalescent and non-infected subjects differ only in innate cytokine production profile. **a–b**, PBMCs in blood collected from a subset of 20 seropositive and 20 seronegative subjects were isolated, analyzed via flow cytometry for cellular composition and stimulated with a SARS-CoV-2 spike peptide mix or vehicle control, or with TLR7 agonist Resiquimod or vehicle control. Activation of CD3^+^CD4^+^, CD3^+^CD8^+^ and CD14^hi^CD16^-^ cells plus cytokine secretion into culture media were measured via flow cytometry. Control corrected (treatment - control) scaled data () is shown in a single clustered heatmap, where scaled anti-spike and anti-RBD IgG titers plus subject age are included (a), or where serpositive subjects are sorted based on time between SARS-CoV-2 positive PCR status and PBMC collection (b). Data was clustered based on Euclidean distances between features (rows) or subjects (columns), with clusters indicated with dendrograms. Serology results are included for each subject (seropositive – purple; seronegative – green) and a time scale is provided. Features most closely associated with antibody titers are highlighted in bold letters.

**Extended Data Figure 4.**
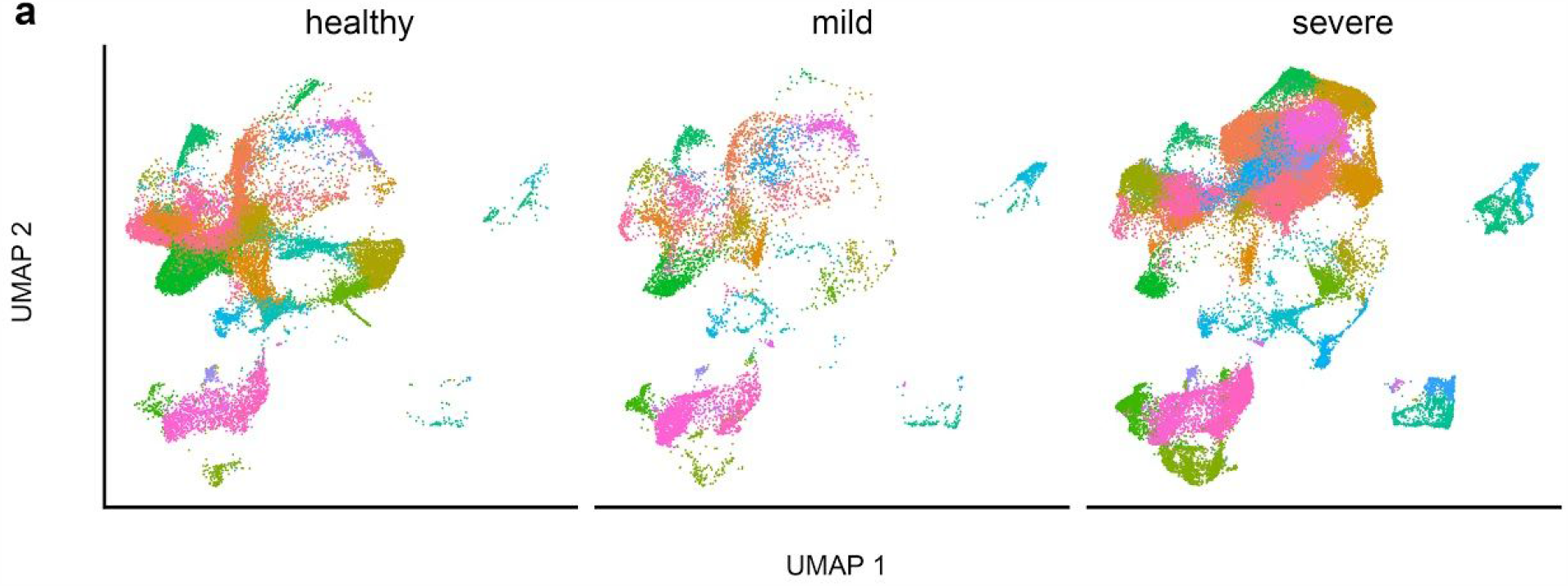
Related to Figure 4: IL-33 production correlates with T cell activation and disease severity SARS-CoV-2 in infected subjects. **a**, Single cell RNA sequencing data from the bronchoalveolar lavage fluid of 3 healthy individual compared to 9 SARS-CoV-2 infected subjects with different disease severities (3 mild; 6 severe) was retrieved from a public database (GSE145926), grouped into cell clusters and shown here by disease status.

**Supplementary Table 1.** Demographics and disease characteristics serology cohorts

| Characteristic | IgG serology cohort | IgM serology cohort |
| --- | --- | --- |
| **Demographics** |  |  |
| No. participants | 155 | 122 |
| Median age (range), years | 36 (5–79) | 34 (5–62) |
| Sex |  |  |
| female - no. (%) | 98 (63.2) | 77 (63.1) |
| male - no. (%) | 57 (36.8) | 45 (36.9) |
| Age |  |  |
| 0-17 | 14 (9) | 13 (10.7) |
| >18 | 141 (91) | 109 (89.3) |
| **Disease characteristics** |  |  |
| Exposure |  |  |
| Professional | 99 (64.5) | 82 (67.2) |
| Household | 44 (28.4) | 30 (24.5) |
| Professional and household | 7 (3.9) | 5 (4.1) |
| Other | 5 (3.2) | 5 (4.1) |
| Symptoms in the last 16 weeks prior serology - no. (%) | 97 (62.6) | 74 (60.7) |
| Median duration of symptoms - days (range) | 10 (1–63) | 11 (1–55) |
| SARS-CoV2 PCR |  |  |
| Positive - no. (%) | 47 (30.3) | 38 (31.1) |
| Negative - no. (%) | 40 (25.8) | 32 (26.2) |
| Not performed - no. (%) | 68 (43.9) | 52 (42.7) |
| Median time between positive PCR and positive serology - days (range) | 49 (10–84) | 45.5 (10–61) |
| **Symptoms** |  |  |
| Cephalgia |  |  |
| Prevalence - no. (%) | 65 (41.9) | 50 (41) |
| Median duration - days (range) | 6 (1–21) | 6 (1–21) |
| Malaise |  |  |
| Prevalence - no. (%) | 63 (40.6%) | 46 (37.7) |
| Median duration - days (range) | 10 (1–63) | 10 (1–48) |
| Cough |  |  |
| Prevalence - no. (%) | 55 (35.5) | 42 (34.4) |
| Median duration - days (range) | 7 (1–21) | 7 (1–20) |
| Fever |  |  |
| Prevalence - no. (%) | 49 (31.6) | 34 (27.9) |
| Median duration - days (range) | 4 (1–45) | 4 (1–14) |
| Musculoskeletal pain |  |  |
| Prevalence - no. (%) | 43 (27.7) | 30 (24.5) |
| Median duration - days (range) | 7 (1–18) | 6.5 (1–18) |
| Anosmia |  |  |
| Prevalence - no. (%) | 40 (25.8) | 29 (23.8) |
| Median duration - days (range) | 7.5 (2–55) | 14 (1–55) |
| Pharyngitis |  |  |
| Prevalence - no. (%) | 37 (23.9) | 31 (25.4) |
| Median duration - days (range) | 3.5 (1–20) | 2.75 (1–14) |
| Dyspnea |  |  |
| Prevalence - no. (%) | 30 (19.4) | 24 (19.7) |
| Median duration - days (range) | 10 (1–21) | 10 (1–21) |
| Weight loss |  |  |
| Prevalence - no. (%) | 24 (15.5) | 17 (13.9) |
| Median value - kg (range) | 3.5 (1–10) | 4.25 (1–10) |
| **Treatment** |  |  |
| Hospitalization | 3 (1.9) | 1 (0.8) |
| Medication |  |  |
| Hydroxychloroquine | 2 (1.3) | 1 (0.8) |
| Antipyretics | 17 (11) | 14 (11.5) |

**Supplementary Table 2.** Demographics and disease characteristics PBMC analysis cohorts

| Characteristic | Seropositive cohort | Seronegative cohort |
| --- | --- | --- |
| **Demographics** |  |  |
| No. participants | 20 | 20 |
| Median age (range), years | 51.5 (14–58) | 37.5 (19–75) |
| Sex |  |  |
| female - no. (%) | 9 (45) | 12 (60) |
| male - no. (%) | 11 (55) | 8 (40) |
| **Disease characteristics** |  |  |
| Median duration of symptoms - days (range) | 15.5 (3–55) | 0 (0–21) |
| SARS-CoV2 PCR |  |  |
| Positive - no. (%) | 18 (90) | 0 (0) |
| Negative - no. (%) | 0 (0) | 8 (40) |
| Not performed - no. (%) | 2 (10) | 12 (60) |
| SARS-CoV2 anti-Spike IgG |  |  |
| Positive - no. (%) | 20 (100) | 0 (0) |
| Negative - no. (%) | 0 (0) | 20 (100) |
| Median time between positive PCR and PBMC collection - days (range) | 66 (29–85) | n.a. |

**Supplementary Table 3.** Key reagents

| Reagent | Company | Identifier |
| --- | --- | --- |
| **Antibodies for ELISA** |  |  |
| Goat anti-Human IgG (H+L) Secondary Antibody, HRP | ThermoFisher Scientific | Cat# 31410 |
| Anti-Human IgM (μ-chain specific)–Peroxidase antibody produced in goat | Sigma | Cat# A0420-1ml |
| **Materials for PBMC isolation** |  |  |
| Lymphoprep | Stem Cell | Cat# 07851 |
| SepMate 50 ml tube | Stem Cell | Cat# 85450 |
| **Virus peptide mixes** |  |  |
| PepMix SARS-CoV2 Spike Glycoprotein | JPT | PM-WCPV-S-1 |
| PepMix HCMVA (pp65) | JPT | PM-PP65-2 |
| **Antibodies for flow cytometry** |  |  |
| AlexaFluor647 anti-human CD3 | Biolegend | Clone: HIT3a<br>Cat# 300322 |
| PE anti-human CD3 | Biolegend | Clone: HIT3a<br>Cat# 300308 |
| BrilliantViolet 711 anti-human CD4 | Biolegend | Clone: OKT4<br>Cat# 317440 |
| FITC anti-human CD4 | Biolegend | Clone: A161A1<br>Cat# 357406 |
| Pacific Blue anti-human CD8 | Biolegend | Clone: HIT8a<br>Cat# 300928 |
| APC anti-human CD8 | Biolegend | Clone: HIT8a<br>Cat# 300812 |
| BrilliantViolet 510 anti-human CD14 | Biolegend | Clone: M5E2<br>Cat# 301842 |
| APC-Cy7 anti-human CD14 | Biolegend | Clone: M5E2<br>Cat# 301820 |
| BrilliantViolet 605 anti-human CD16 | Biolegend | Clone: 3G8<br>Cat# 302039 |
| APC-Cy7 anti-human CD16 | Biolegend | Clone: 3G8<br>Cat# 302017 |
| PE anti-human CD19 | Biolegend | Clone: HIB19<br>Cat# 302208 |
| APC-Cy7 anti-human CD19 | Biolegend | Clone: HIB19<br>Cat# 302217 |
| PE/Dazzle 594 anti-human CD27 | Biolegend | Clone: M-T271<br>Cat# 356421 |
| PE-Cy7 anti-human CD45RA | Biolegend | Clone: HI100<br>Cat# 304112 |
| APC-Cy7 anti-human CD56 | Biolegend | Clone: HCD56<br>Cat# 318332 |
| AlexaFluor 488 anti-human CD62L | Biolegend | Clone: DREG56<br>Cat# 304816 |
| BrilliantViolet 785 anti-human CD69 | Biolegend | Clone: FN50<br>Cat# 310932 |
| PE/Dazzle 594 anti-human CD134 | Biolegend | Clone: ACT35<br>Cat# 350019 |
| BrilliantViolet 785 anti-human CD197 | Biolegend | Clone: G043H7<br>Cat# 353230 |
| AlexaFluor 488 anti-human FOXP3 | Biolegend | Clone: 206D<br>Cat# 320111 |
| AlexaFluor647 anti-human IFNγ | Biolegend | Clone: 4SB3<br>Cat# 502516 |
| AlexaFluor700 anti-human TNF | Biolegend | Clone: MAb11<br>Cat# 502928 |
| PE-Cy7 anti-human TNF | eBioscience | Clone: TN3-19.12<br>Cat# 25-7423-82 |
| FITC anti-human IL-1 beta | eBioscience | Clone: CRM56<br>Cat# 11-7018-42 |
| eFluor 450 anti-human IL-6 | eBioscience | Clone: MQ2-13A5<br>Cat# 48-7069-42 |
| APC anti-human IL-10 | Biolegend | Clone: JES3-9D7<br>Cat# 501409 |
| **Other reagents for flow cytometry** |  |  |
| Live/Dead Fixable Aqua Dead Cell Stain Kit | ThermoFisher Scientific | Cat# 34957 |
| Live/Dead Fixable Blue Dead Cell Stain Kit | ThermoFisher Scientific | Cat# L34962 |
| Purified NA/LE Human BD Fc block | BD | Cat# 564765 |
| **Commercial kits** |  |  |
| TMB substrate set | Biolegend | Cat# 421101 |
| Foxp3/Transcription factor staining buffer set | eBioscience | Cat# 00-5523-00 |
| Fixation/Permeabilization Solution Kit | BD | Cat# 554714 |
| LEGENDplex Human Inflammation Panel 1 | Biolegend | Cat# 740809 |

